# Thrombotic and thromboembolic events, with or without thrombocytopenia, following viral vector-based COVID-19 vaccines administration: a systematic review protocol

**DOI:** 10.1101/2024.02.22.24303207

**Authors:** José Ramos-Rojas, Javiera Peña, Carlos Pinto-Díaz, Valentina Veloso, Gabriel Rada, Helvert Felipe Molina-León

## Abstract

**Background:** Viral vector-based COVID-19 vaccines have proven to be effective and safe in clinical trials and post-authorization studies. Although infrequent, some serious thrombotic and thromboembolic events following immunization have emerged, and causality assessment committees must consider and critically assess different sources of evidence to inform their decisions about whether these events supposedly attributable to vaccination or immunization (ESAVI) are associated with the vaccine or are coincidental. Therefore, this systematic review aims to gather information on the association and biological mechanisms between thrombotic and thromboembolic events, with or without thrombocytopenia, and the administration of viral vector-based COVID-19 vaccines.

**Methods:** We will conduct a systematic review following the evidence synthesis framework proposed by the Pan American Health Organization to support the ESAVI causality assessment. We will search for primary clinical and preclinical studies in the Epistemonikos’ COVID-19 L.OVE (Living Overview of the Evidence) repository, a comprehensive and validated source of COVID-19 evidence. We will include studies reporting any thrombotic or thromboembolic event, with or without thrombocytopenia, after the administration of a viral vector-based COVID-19 vaccine. The screening and data extraction will be performed by two independent authors. We will assess the risk of bias by two reviewers using the appropriate tool for each study design. Discrepancies will be discussed or resolved by a third author. We will use GRADE to assess the certainty of evidence for clinical studies and prepare summary of findings tables. For individual-based (case series and case reports) and preclinical studies, we will summarize the results in descriptive tables.

**Expected results and implications:** This will be the first systematic review using the evidence synthesis framework for ESAVI causality assessment, currently under validation by the Pan American Health Organization and the Epistemonikos Foundation. By gathering clinical and preclinical evidence, it is expected to inform about the risks of thromboembolic events following vaccination with viral vector-based COVID-19 vaccines, and also the possible underlying biological mechanisms. Policymakers, such as safe vaccination committees, and other evidence synthesis authors could replicate this novel methodology to strengthen the evidence-based ESAVI causality assessment.

## Introduction

During the COVID-19 pandemic, an unprecedented effort in developing healthcare technologies emerged. Various anti-SARS-CoV-2 vaccines were developed during this period, reducing the morbidity and mortality associated with COVID-19 through different mechanisms of action or platforms, one of them being viral vector vaccines (1). These vaccines use genetically engineered viral vectors to introduce key antigens of pathogens into human cells (2). This induces an immune response but has no pathogenicity (3).

As of 2022, over 25 vaccine candidates have been identified in clinical trials considering human adenoviruses and chimpanzee adenovirus as viral vectors (4). Among them, both Ad26.COV2.S and AZD1222 (ChAdOx1 nCoV-19), developed by Janssen and AstraZeneca, respectively, were the most extensively used. Both vaccines belong to the non-replicating category, demonstrating its effectiveness and safety after intramuscular administration in large clinical trials since 2020, collectively including over 70,000 participants (4).

The authorization, widespread distribution and subsequent post-authorization studies confirmed its effectiveness and contributed to the safety monitoring at large-scale observational studies, with low rates of reported serious adverse events (3). However, even though they are infrequent, the potential occurrence of these adverse events must be consistently considered and analyzed through vigilance systems to ensure the safety of vaccines, the population’s health, and to provide accurate information for public awareness (5).

One of the adverse events identified as a potential consequence of the administration of viral vector-based COVID-19 vaccines is thrombosis, including thrombosis events with thrombocytopenia. Many of these cases have been associated with autoantibodies against the platelet factor 4 (PF-4) antigen, resembling those observed in patients with autoimmune heparin-induced thrombocytopenia (1). Its occurrence is very rare, with reported incidences of less than 1 per 100,000 vaccinated persons (6) and could have severe health consequences for individuals and potentially be fatal (7).

However, its manifestation may be linked to other causes (6). It has been observed that over 90% of the reported cases of pulmonary thromboembolism following vaccination have other relevant risk factors (7). For instance, COVID-19 infection is a well-known risk factor for thrombosis development, so its occurrence should be carefully analyzed to determine whether there is a causal association with vaccination or if it is merely coincidental (6). For this reason, the analysis of cases of thromboembolic events or thrombosis with thrombocytopenia following vaccine administration must be studied diligently.

Conducting a causality assessment of events supposedly attributable to vaccination and immunization (ESAVI) is highly relevant from various perspectives. Firstly, understanding potential vaccine complications is crucial for effective treatment and risk-benefit assessments in mass distribution. Secondly, the reported severe complications may fuel vaccine hesitancy, exacerbating pre-existing distrust. Thirdly, given that storage conditions are simpler for this type of vaccine, they are more likely to be used in developing countries than other alternatives (for example mRNA vaccines). Therefore, it becomes even more necessary to minimize any risk of associated adverse events (1).

For this causality assessment, it is essential to consider various levels of analysis and diverse sets of evidence. Therefore, the objective of this review is to gather information on the association and biological mechanisms between thrombotic and thromboembolic events, with or without thrombocytopenia, and the administration of viral vector-based COVID-19 vaccines.

Additionally, it aims to support the formulation of individual-level causality conclusions for causality assessment committees through an evidence-based health care approach. This protocol is the first one conducted following the guidelines of a novel evidence synthesis framework related to ESAVI, contributing to the ongoing causality discussion led by the Pan American Health Organization and the Epistemonikos Foundation.

## Methods

We will conduct a systematic review following the evidence synthesis framework proposed by the Pan American Health Organization to support the ESAVI causality assessment. This protocol adheres to the PRISMA-P (*Preferred Reporting Items for Systematic Review and Meta-Analysis Protocols*) checklist (8).

### Research questions

1. Can the administration of viral vector-based COVID-19 vaccines increase the risk of developing thrombotic and thromboembolic events, with or without thrombocytopenia?
2. Are there known biological mechanisms that support a possible causal association between viral vector-based COVID-19 vaccines and the development of thrombotic and thromboembolic events, with or without thrombocytopenia?

### Information sources

We will conduct searches in the Epistemonikos’ COVID-19 L.OVE repository, that is continuously updated through searches in different databases, trial registries, and preprint servers (9). This repository has been validated as a complete and comprehensive source of COVID-19 related articles (9–11). We will not apply language or date restrictions. The search strategy is available in the <u>supplemental file 1</u>.

### Eligibility criteria

#### a) Participants

We will include clinical studies in individuals at risk of developing thrombosis and thromboembolism, with or without thrombocytopenia, as well as preclinical studies in humans, and animal or *in vitro* models.

We will include studies reporting thrombotic and thromboembolic events confirmed by imaging, surgical procedures, or pathological examination by autopsy or biopsy. Imaging studies will consider: ultrasound (compression with or without doppler),

Within imaging studies, we will consider ultrasound (compression with or without Doppler), contrast enhanced computed tomography, contrast venography, magnetic resonance angiogram, echocardiogram, ventilation/perfusion scintigraphy, conventional angiography and digital subtraction angiography. Additionally, we will include studies informing procedures that confirm the presence of a thrombus (for example, thrombectomy) and pathological findings (biopsy or autopsy) consistent with thrombosis or thromboembolism (7).

In cases related to thrombosis and thrombocytopenia, we will include those informing at least a platelet count lower than 150 × 10^9/L, a platelet count less than the lower limit of the local laboratory values, or a ≥50% decrease compared to a previous count. We will also consider the persistence of severe cephalea within 5 days after the vaccination with a D-dimer elevation superior to 8 x ULN (corresponding to >4000 μg/ml (FEU)). In addition, we will consider a risk window of 4 to 30 days of symptom initiation after vaccination; up to day 42 for deep vein thrombosis or pulmonary embolisms.

For the individual-based studies, specially case series and case reports of thrombotic and thromboembolic events, we will exclude those in which pathological or imaging findings consistent with said conditions are not reported.

#### b) Intervention

We will include studies evaluating the effect of the administration of any viral vector-based COVID-19 vaccine (Ad5-nCoV, Ad26.COV2-S, ChAdOx1-S and rAd26-S+rAd5-S). We will also include studies evaluating the effects of the administration of specific viral vector vaccines components in preclinical or *in vitro* models. Studies not specifying the type of vaccine administered and those in which the presence of defects in the administered vaccine or incidents in the vaccination process will be excluded.

#### c) Comparison

We will include studies comparing the intervention with non-vaccinated groups, who received a placebo, other COVID-19 vaccine platforms, or a non-exposed period. We will also include non-comparative studies such as case reports and case series or descriptive observational studies.

#### d) Outcomes

We will include studies reporting the development of thrombotic and thromboembolic events, with or without thrombocytopenia. Specifically, studies reporting events of cerebral venous thrombosis, cerebral venous sinus or jugular vein thrombosis, deep vein thrombosis, pulmonary embolisms, intra-abdominal venous thrombosis (hepatic, splanchnic, mesenteric, and/or renal), cerebral ischemia, myocardial infarction due to coronary artery thrombosis, limb ischemic events, and other ischemic events in unusual sites.

From observational descriptive studies, outcomes of interest are the background rates of the adverse events at a population level (i.e., before the administration of COVID-19 vaccines). In individual-based studies (case reports and case series), outcomes of interest are the demographic and clinical characteristics of the cases, the risk window or period until the onset of the adverse event, available laboratory tests, description of the clinical evolution, treatment administered, and history of previous adverse events per similar exposure (rechallenge).

We will exclude studies reporting thrombocytopenia without thrombotic or thromboembolic events.

#### e) Study design

We will include primary studies of any design. Review articles (of any kind) will be excluded.

### Selection process

Study selection will be performed in Collaboratron, the Espitemonikos’ screening software (12) by six review authors who will screen by title and abstract (JT, JP, MB, VV, FC and SF). Then, we will examine full texts to determine if they fulfill the eligibility criteria. In both processes each reference will be examined by duplicate and we will resolve discrepancies by discussion or by a third author. We will report the selection process as a PRISMA flowchart. We will separately inform the source of evidence identified in the following categories: a) clinical studies (experimental and observational comparative), b) preclinical studies, and c) individual-level studies (case reports and case series). The list of excluded studies by full-text and the reasons for exclusion will be reported in the supplementary material of the systematic review.

### Data collection

We will extract data from the primary studies by duplicate (i.e., two independent authors: JP, JT, MB and SF) using standardized forms designed for this project. We will perform a preliminary pilot stage and resolve discrepancies by consensus. We will collect the following data according to the study design:

- For all included studies:
  - Author, year, title, country and journal.
  - General study characteristics: intervention/exposure assignment, type of comparison and follow-up temporality (cross-sectional, prospective or retrospective).
  - Adverse event characteristics: type of thrombotic event and presence of thrombocytopenia. Additionally, if the diagnostic Brighton criteria, International Classification of Diseases - ICD-, or any other formal case definition was used for identifying cases of these ESAVI.
- Experimental and observational studies:
  - Participant characteristics: age, sex, previous COVID-19 infection, pregnancy or breastfeeding status, risk factors (tobacco use, hormone replacement therapy, use of oral contraceptives, immobility, and recent trauma or surgery), comorbidities (obesity, cancer, congenital cardiac diseases, atrial fibrillation, sickle cell disease), medication use (heparin, immunoglobulins, antidepressants and/or antipsychotics) and concomitant administration of other COVID-19 vaccines or regular vaccination schedules (for example, varicella zoster vaccine, influenza vaccine, human papillomavirus vaccine, antipneumococcal vaccine, tetanus vaccine, diphtheria-tetanus-pertussis vaccine).
  - Intervention characteristics: type of administered vaccine, previous COVID-19 vaccination schedule, outcome measurement method.
  - Comparison characteristics: placebo administration, non-vaccinated group, or non-exposed period. If a different COVID-19 vaccine is administered, we will extract data about the type of vaccine and dosing.
  - Outcomes: number and incidence of the adverse events of interest, estimates of effects as risk estimates, incidence rates ratios, hazard ratios, or observed/expected rates. If available, we will extract the adjusted estimates and record the adjustment variables. We will also collect the time interval or risk window (as days) between vaccination and ESAVI onset.
- Individual-level studies (case reports and case series):
  - Participant characteristics: age, sex, comorbidities (e.g., obesity, cancer, inflammatory bowel diseases, migraine, obstructive sleep apnea, vasculitis, congenital cardiac diseases, atrial fibrillation, sickle cell disease), pregnancy, drugs affecting coagulation (e.g., heparin).
  - Intervention characteristics: administered vaccine and dosing, report of administration errors or vaccine defects.
  - Outcome characteristics. For thrombotic and thromboembolic events: risk window (measured as days), clinical characteristics of the adverse event, laboratory findings (D-dimer, fibrinogen, anti-PF4 antibodies measured by ELISA or by functional/platelet activation assays), imaging studies, administered treatment, clinical evolution, alternative cause study and differential diagnoses. For thrombotic events including thrombocytopenia: platelet count, specifying the method used and the upper and lower values of the local laboratory. For all events: history of previous exposition (positive rechallenge).
- Preclinical studies (animal or *in vitro*) and in humans reporting biological mechanism evidence:
  - Study design characteristics: randomization and adverse event induction method.
  - Population characteristics. For animals or *in vitro* studies, we will extract the model used. For studies in humans, we will extract data of studied participants (age, sex, comorbidities and type of biological sample).
  - Intervention characteristics: administered vaccine, dosing and administration route.
  - Outcomes: changes in hematological profiles, immunological response (antibodies), inflammatory markers and other biomarkers related to the development of thrombotic and thromboembolic events, with or without thrombocytopenia.

### Risk of bias assessment

For randomized trials, we will assess the risk of bias using the RoB-2 tool, developed by Cochrane (13). For non-randomized studies (experimental and observational comparative), we will assess the risk of bias using the ROBINS-I tool, developed by Cochrane (14).

For individual-based studies (case reports and case series), we will assess their reporting quality using the Joanna Briggs Institute’s checklist for case reports (15).

For preclinical studies, we will use the SYRCLE risk of bias tool, adapted from the RoB tool for these types of studies (16)..

### Data synthesis

Firstly, to summarize the findings from different sources of evidence, we will classify the included studies using the Design Algorithm for Medical Literature on Intervention proposed by Seo et al. (2015) (17). This will allow us to standardize the classification of studies according to their design and select the proper critical assessment tools.

After that, considering the type of study and reported outcomes, we will classify the studies into three sets of evidence:

- Epidemiological evidence: clinical studies that allow us to estimate risks and associations.
- Preclinical evidence: preclinical studies (*in vivo* or *in vitro*) that contribute to the understanding of biological mechanisms involved in the development of the adverse event. We will also consider studies in humans that inform on the biological mechanism evidence.
- Individual-level evidence: studies in humans (mostly case reports and case series) that provide suggestive elements of possible biological mechanisms involved in the development of the adverse event.

Once classified, we will perform a narrative synthesis of the included studies. We will report the number and type of study identified per evidence set, number of participants, and characteristics of the participants, intervention and comparison.

For the epidemiological evidence set, if data from risks, associations, incidence rates (estimates and dispersion measures), or basal rates of the adverse events of interest are reported, and if there is no clinical and methodological heterogeneity, we will perform a quantitative synthesis or meta-analysis. If there is not enough data for quantitative synthesis, or there is considerable heterogeneity-clinical or methodological-we will narratively summarize the findings (i.e., the number of participants or administered vaccines per each group and the reported effect estimates with their confidence intervals).

For preclinical studies, we will narratively describe the outcomes reported in the included studies. If the reported outcomes are comparable and there is sufficient data, we will perform a quantitative synthesis or meta-analysis. However, frequently in this type of studies, there is high heterogeneity and usually there is insufficient reported data.

For individual-level studies (case reports and case series) we will perform a narrative synthesis of the main findings.

### Certainty of evidence assessment

We will assess the certainty of evidence for the clinical and preclinical studies. For clinical studies of the epidemiological set, the certainty of evidence will be assessed using the GRADE (Grading of Recommendations Assessment, Development and Evaluation) approach (18), which assesses the confidence in the effect estimates evaluating different factors that can reduce the quality of evidence (i.e., study design and risk of bias in the included studies, inconsistency, indirectness, imprecision, and publication bias) and others that increases the quality of the evidence (i.e., a large magnitude of effect, confounding factors, dose-response gradient). In the context of this ESAVI systematic review, according to the proposed PAHO framework (currently under development), we will start the evaluation of non-randomized trials from a moderate level of confidence.

For preclinical studies, we will assess the certainty of evidence following the proposed methods of the SYRCLE (Systematic Review Centre for Laboratory Animal Experimentation) working group, which applies the GRADE approach with some considerations to be used in the context of these types of studies (12).

Finally, regarding the individual-level studies, we will not evaluate the certainty of evidence, since their use is mainly focused on narratively describing relevant elements for the causality discussion.

We will prepare summary of findings (SoF) tables for the epidemiological and preclinical sets (13). As for individual-level studies, we will elaborate a descriptive table of the main findings and their quality of report, as suggested in the evidence synthesis framework for the ESAVI causality assessment developed by PAHO.

## Expected results and implications for ESAVI causality assessment

This systematic review will be the first to use the reference manual for evidence synthesis related to ESAVI proposed by PAHO and the Epistemonikos Foundation which is currently in their validation stage. In this regard, performing this systematic review is essential to evaluate the applicability of the methods framework proposed in the manual.

Additionally, the results of this review may be used by regional, national or subnational safe vaccination committees to support the causality discussion of individual cases of thrombosis and thromboembolism, with or without thrombocytopenia, following the administration of viral vector vaccines against COVID-19.

As far as we know, this is the first review integrating both epidemiological and biological mechanisms evidence to inform about the risks of thrombotic and thromboembolic events after vaccination with viral vector COVID-19 vaccines. Hence, the methods reported in this protocol can be replicated by other authors and be used to strengthen the evidence-informed ESAVI causality assessment process.

## Supporting information

Supplemental table 1. Search strategy.

## Data Availability

All data produced in the present study are available upon reasonable request to the authors.

## Acknowledgements

We would like to acknowledge Francy Cantor, Magdalena Bignon, Sebastián Pinto, and Sergio Fernández from the Epistemonikos methods team for their contribution in the preliminary screening of this systematic review.

