## Supplemental table 1. Search strategy. for "Thrombotic and thromboembolic events, with or without thrombocytopenia, following viral vector-based COVID-19 vaccines administration: a systematic review protocol"

#### Appendix 1. Search strategy for thrombotic and thromboembolic events following viral vector-based COVID-19 vaccines in the Epistemonikos’ COVID-19 L.OVE.

| **Component** | **#** | **Boolean strategy** |
| --- | --- | --- |
| **Intervención** | | |
| Viral vector COVID-19 vaccines | #1 | dendritic* OR aivita |
|  | #2 | sars* OR covid* |
|  | #3 | vaccine |
|  | #4 | #1 AND #2 AND #3 |
|  | #5 | Influenza OR flu OR "flu-based" |
|  | #6 | vector AND vaccine |
|  | #7 | #5 AND #23 AND #6 |
|  | #8 | Merck OR MSD |
|  | #9 | covid* OR SARS* |
|  | #10 | vaccine |
|  | #11 | #8 AND #9 AND #10 |
|  | #12 | janssen* OR johnson* OR "j&J" OR "j & J" |
|  | #13 | vaccin* |
|  | #14 | coronavir* OR "corona virus" OR "virus corona" OR "corono virus" OR covid* OR "2019-ncov" OR cv19* OR "cv-19" OR "cv 19" OR "n-cov" OR ncov* OR antisars* OR "anti-sars-cov-2" OR "anti-sars-cov2" OR "anti-sarscov-2" OR "anti-sarscov-2" |
|  | #15 | #31 OR #14 |
|  | #16 | #12 AND #13 AND #15 |
|  | #17 | serum AND institute AND India |
|  | #18 | Oxford* OR Astra* OR "SK Bio" |
|  | #19 | #17 OR #18 |
|  | #20 | vaccin* |
|  | #21 | #19 AND #20 |
|  | #22 | Serum AND Institute AND India AND vaccin* |
|  | #23 | COVID* OR SARS* |
|  | #24 | gamaleya* AND vaccine* |
|  | #25 | #23 AND #24 |
|  | #26 | Sputnik* AND Light |
|  | #27 | Cansino OR AD5* OR "type 5" |
|  | #28 | #27 AND #29 |
|  | #29 | vaccin* OR immunization OR immunogenic OR immunisation OR reactogenic* |
|  | #30 | wuhan* |
|  | #31 | #30 AND #33 |
|  | #32 | covid* |
|  | #33 | virus OR viruses OR viral |
|  | #34 | #32 AND #33 |
|  | #35 | "covid-19" OR covid19* OR "covid 19" OR "2019-nCoV" OR cv19* OR "cv-19" OR "cv 19" OR "n-cov" OR ncov* OR "sars-cov-2" OR "sars-cov2" OR "2019-ncov" OR "covid-19-related" OR "SARS-CoV-2-related" OR "SARS-CoV2-related" OR "2019-nCoV-related" OR "cv-19-related" OR "n-cov-related" |
|  | #36 | #31 OR #34 OR #35 |
|  | #37 | #29 AND #36 |
|  | #38 | COH04S1* OR "MVA-SARS-2-S" OR "AV-COVID-19" OR DelNS1* OR V591* OR "V-591" OR V590* OR "V-590" OR Ad26* OR "jnj-78436735" OR "jnj 78436735" OR jnj78436735* OR VAC31518* OR "VAC-31518" OR "VAC-31518" OR Jcovden* OR ChAdOx1* OR AZD1222* OR "AZD 1222" OR "AZD-1222" OR Covishield* OR Vaxzevria* OR covishield* OR "Gam-COVID-Vac" OR "Gam COVID Vac" OR GamCOVIDVac* OR sputnik* OR "GRAd-COV2" OR "AD5-nCOV" OR "AD5 nCOV" OR AD5nCOV* OR Convidecia* OR PakVac* OR TMV-083* OR "TMV 083" OR TMV083* |
|  | #39 | #4 OR #7 OR #11 OR #11 OR #16 OR #21 OR #22 OR #25 OR #26 OR #28 OR #37 OR #38 |
| **Population** | | |
| Thrombocytopenia | #40 | thrombocytop* OR thrombopen* OR "low platelet" OR "low platelets" |
| Thrombosis | #41 | thrombos* OR thrombot* OR embol* OR thromboemboli* |
| **Total** | | |
| #39 AND (#40 OR #41) | | |
